# Predicting Emerging Themes in Rapidly Expanding COVID-19 Literature with Dynamic Word Embedding Networks and Machine Learning

**DOI:** 10.1101/2021.01.14.21249855

**Authors:** Ridam Pal, Harshita Chopra, Raghav Awasthi, Harsh Bandhey, Aditya Nagori, Amogh Gulati, Ponnurangam Kumaraguru, Tavpritesh Sethi

**Affiliations:** Indraprastha Institute of Information Technology Delhi, India; All India Institute of Medical Sciences, New Delhi, India; CSIR-Institute of Genomics and Integrative Biology, Delhi, India; Maharaja Surajmal Institute of Technology, GGSIPU, New Delhi, India

## Abstract

Evidence from peer-reviewed literature is the cornerstone for designing responses to global threats such as COVID-19. The collection of knowledge and interpretation in publications needs to be distilled into evidence by leveraging natural language in ways beyond standard meta-analysis. Several studies have focused on mining evidence from text using natural language processing, and have focused on a handful of diseases. Here we show that new knowledge can be captured, tracked and predicted using the evolution of unsupervised word embeddings and machine learning. Our approach to decipher the flow of latent knowledge in time-varying networks of word-vectors captured thromboembolic complications as an emerging theme in more than 77,000 peer-reviewed publications and more than 11,000 WHO vetted preprints on COVID-19. Furthermore, machine learning based prediction of emerging links in the networks reveals autoimmune diseases, multisystem inflammatory syndrome and neurological complications as a dominant research theme in COVID-19 publications starting March 2021.

## Introduction

COVID-19 pandemic continues to be an enigma with its diverse clinical presentation, controversial evidence for treatment, fast-tracked vaccine development and unclear systemic implications. More than 200 countries have been affected by the pandemic with around 75 million confirmed cases and more than 1.6 million deaths recorded till 22nd December 2020 (1). The literature around COVID-19 is growing at an exponential rate with more than 77,000 peer-reviewed research articles and more than 11,000 WHO vetted preprints(2). Understanding COVID-19 in the context of evolving themes is important as knowledge synthesis from peer-reviewed literature becomes increasingly difficult for researchers, clinicians, and policymakers alike. Methods such as topic modeling and sentiment analysis have been previously carried out comparing pre-print with peer-reviewed literature only over a short time span (3). Our paper aims to fill these gaps through *EvidenceFlow*, a publicly available interactive dashboard for tracking literature trends using alluvial diagrams, projection of influential entities, and network analysis across different months. We are reporting for the first time, the use of unsupervised word embeddings, link prediction in dynamic networks of entities and machine learning to predict emerging themes in COVID-19 literature and making these publicly available as a web application. The current word embedding models were trained upon the abstracts obtained from the WHO Database and will be updated with new publications and pre-prints as these become available on a monthly basis.

The abstract of articles holds a substantial amount of information about the literature. Named entities play a crucial role in deducing valuable information from large amounts of text and influencing the trends of literature (4). Models pre-trained on biomedical, scientific and clinical benchmark datasets, can be used for extraction of a variety of clinical entities such as diseases, symptoms, chemicals and adverse drug reactions from continuous text. The relative context of these entities changes over time leading to a ‘semantic shift’ in similarity with other words (5). Unsupervised word embeddings have been used to capture complex science concepts using the semantic relationship signified by cosine similarity (6). We studied the evolution of literature based on changing cosine similarity between extracted entities in weighted temporal networks.

Link Prediction has been defined as the task of predicting the existence of links between two nodes in a complex network based on a set of topological features. The problem of link prediction in real-world temporal networks has been explored a lot in recent years (7), primarily in online social media networks where nodes are represented by users and edges by the relationship between them. Supervised learning methods based on topological proximity measures have been vastly used to capture the temporal shift of links within networks (8,9). We forecasted semantic and topological proximity features of entity pairs based on their temporal trends in previous months and used these as features to predict links between clinical entities extracted from textual data over T time intervals. Given the vast research found on online social networks, our framework differs from the standard link prediction models as it studies the concept by applying named entity recognition in the scientific literature. The predicted links were used to create a network weighted by forecasted cosine similarity for detecting communities of entities that tend to reflect on themes of the articles published in that month. The schematic representation of workflow has been demonstrated (Figure 1).

**Figure 1:**
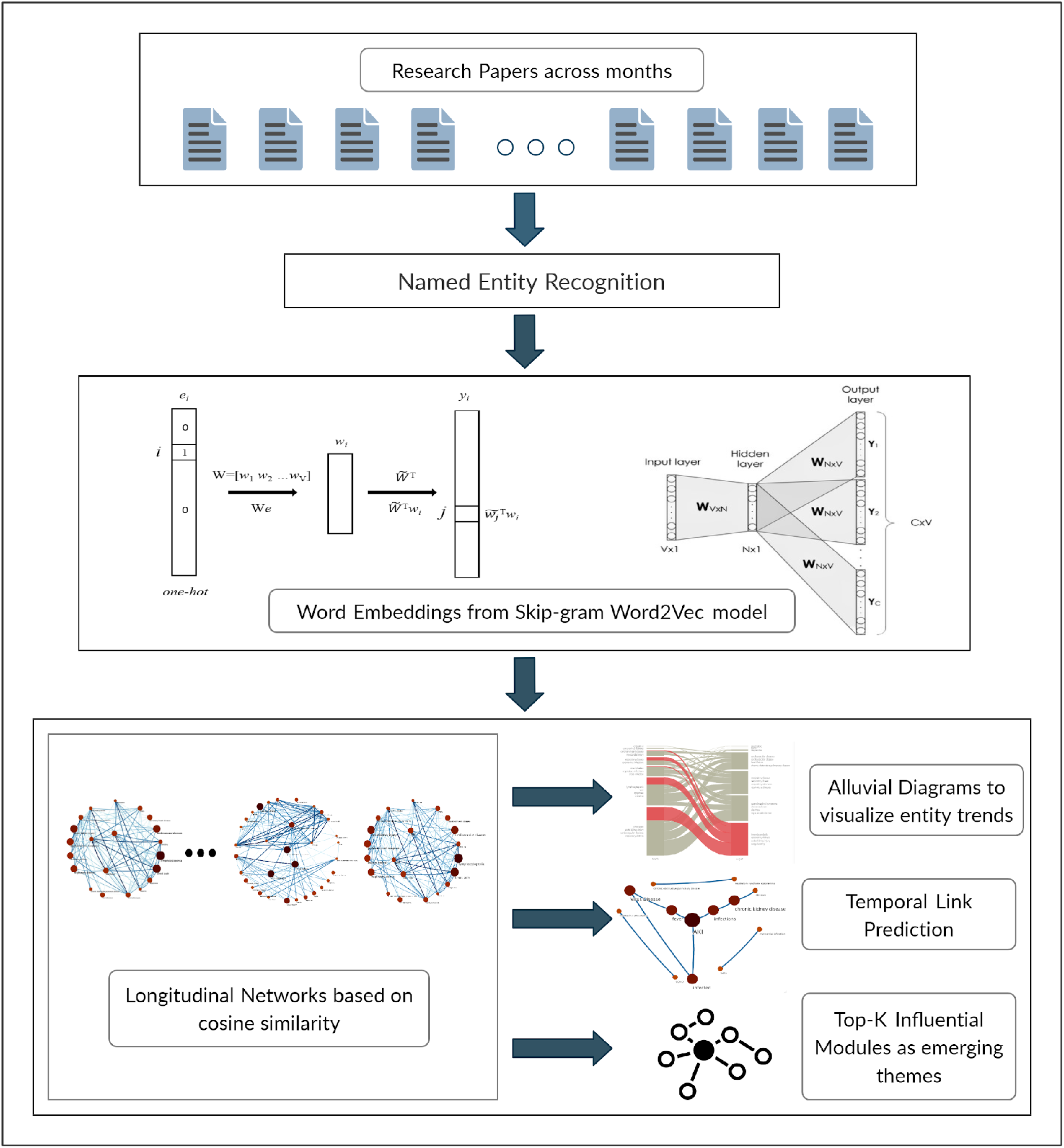
Graphical representation of proposed framework explaining the complete workflow. The pipeline takes abstracts as input from which entities are extracted using NER. Embeddings are generated which are used as features for longitudinal networks. These networks are used for visualizing the trends using alluvial diagrams, temporal link prediction and predicting top-k influential modules for theme prediction.

### Implementation and Availability

*EvidenceFlow*, our web application with results of online tracking and prediction of emerging themes is available publicly at https://evidenceflow.tavlab.iiitd.edu.in/.

## Results

A total of 29937 distinct diseases and 26087 distinct chemicals were identified and top entities are shown in Supplementary Figure 1 (a) and (b). The latent space of word embeddings around the keyword ‘COVID-19 disease’ was also visualized in Supplementary Figure 1 (c).

We examined an approach to graphically explore the temporal trends in the literature using alluvial diagrams based on dynamic and homogeneous networks of prevalent medical entities and their associated cosine similarities. A detailed inference of the alluvial diagram across the months of March and August in Figure 2(b) depicted the emergence of thromboembolic complications as the most important module. In March, the dominant modules were chest pain, acute kidney injury and lymphocytopenia depicting lesser traces of thromboembolic complications in the initial months. The network of August as shown in Figure 2(a) and Supplementary Figure 2 evidently captures the rising influence of nodes linked to thromboembolism, gastrointestinal symptoms, respiratory and cardiovascular diseases.

**Figure 2:**
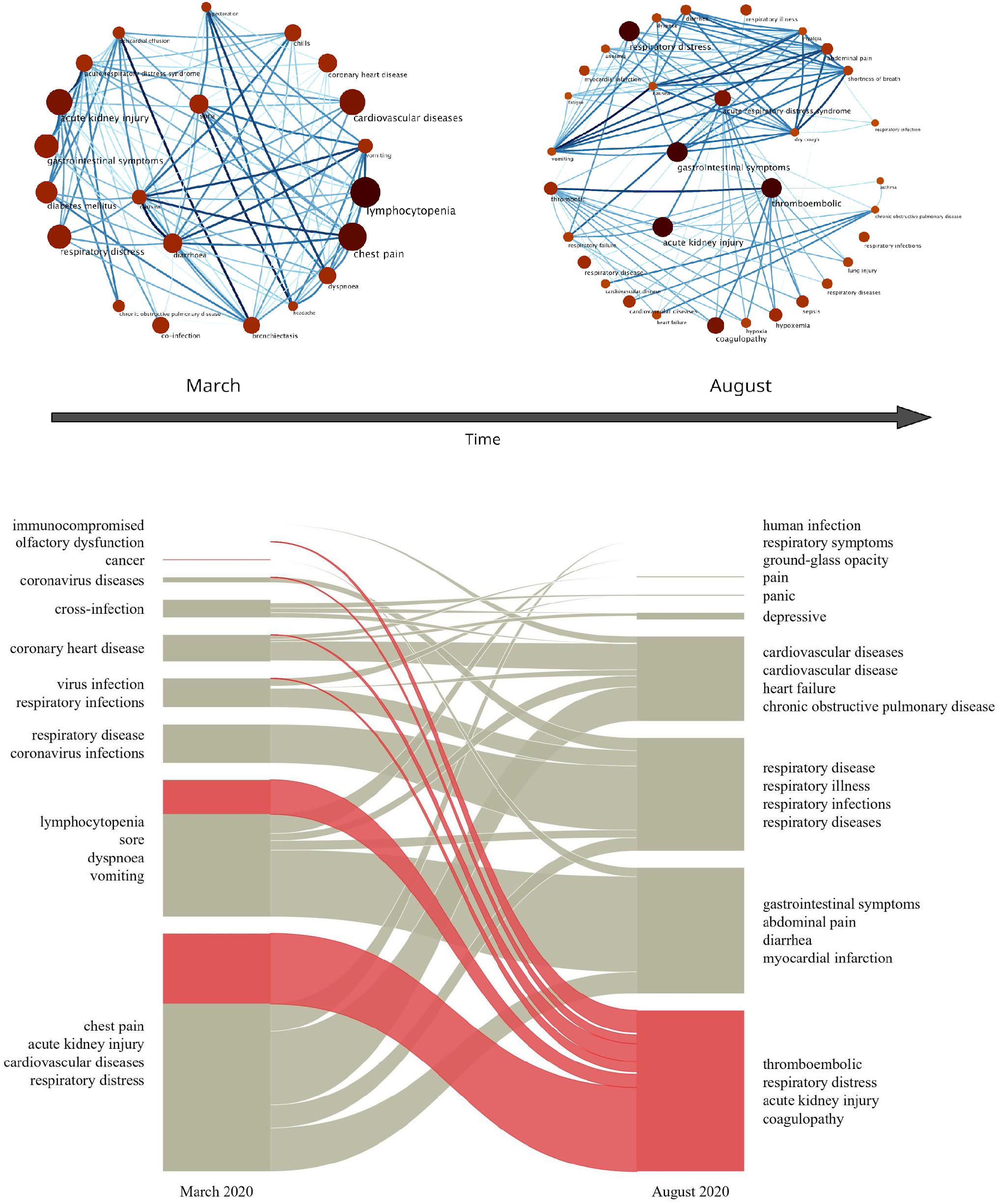
a) Longitudinal source network from March 2020 to August 2020. (b) Alluvial diagram for tracking the trends from March 2020 to August 2020. The alluvial diagram eases tracing the trends of temporal dynamics of literature across different months. The diagram clearly illustrates the emergence of thromboembolic complications as a major theme by August.

We further advance the analysis of trends to predicting links between entity pairs for the upcoming months. Our proposed framework for Temporal Link Prediction effectively forecasted five proximity scores, including semantic and topological measures, between node pairs by modeling its time series using the ARIMA model. The associations between diseases for the successive month were predicted as links using supervised learning, on the basis of dynamic networks belonging to the previous months. Our results show that among the five classifiers (Supplementary Table 1), the Random Forest model classified links with a mean AUC ROC score of 0.77 in the test set (Figure 3).

**Figure 3:**
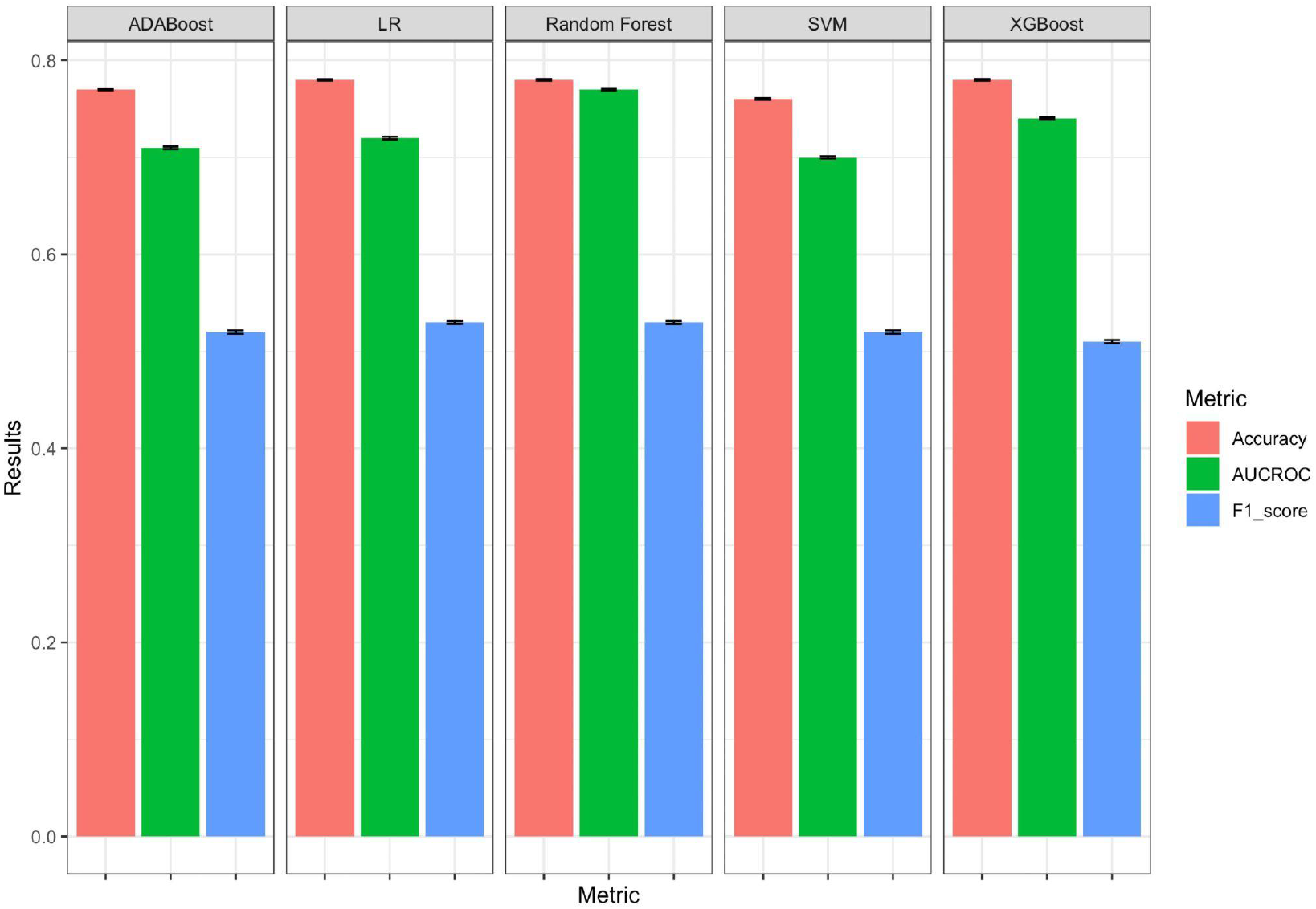
Results of temporal link prediction between entities for the month of February 2021, with a margin of error for 95% Confidence Intervals. The mean value of metrics has been recorded by testing the models on a resampled test set.

The predicted and original network of February 2021 were clustered into modules using the Infomap algorithm. The intersection of nodes between respective modules was analyzed for validating the effectiveness of the proposed framework. Five modules are mentioned in Table 1 along with their respective IOU and a subset of intersecting entities, which have been interpreted to represent broad themes. Psychological conditions as a theme was detected with the highest score of IOU. Supplementary Figure 5 shows the percentage of articles mentioning entities from each module respectively. Respiratory infections (Module 4) evidently remained the most discussed theme, followed by adverse events (Module 1) and comorbidities (Module 2).

**Table 1:** Results of community detection on predicted network of February 2021. IOU denotes the Intersection Over Union of the nodes present in the predicted and original network’s module. A subset of intersecting nodes is mentioned which collectively signify a theme.

| Predicted network | Original network | IOU | Theme: Subset of Intersecting nodes |
| --- | --- | --- | --- |
| Module 1 | Module 1 | 0.61 | Adverse events:<br>respiratory failure, pulmonary embolism, acute kidney injury, hyperinflammation, sepsis |
| Module 2 | Module 3 | 0.58 | Comorbidities:<br>obesity, breast cancer, diabetes, hypertension, asthma, dementia, chronic kidney disease |
| Module 3 | Module 4 | 0.41 | Symptoms:<br>anosmia, shortness of breath, fever, fatigue, diarrhea, myalgia, dyspnea, headache, cough |
| Module 4 | Module 2 | 0.55 | Respiratory illness:<br>MERS, acute respiratory syndrome coronavirus 2, respiratory infections |
| Module 6 | Module 5 | 0.72 | Psychological conditions:<br>PTSD, psychological distress, loneliness, burnout, anxiety, depression, traumatic |

## Discussion

We developed an approach to take advantage of dynamic and homogeneous networks of medical entities and their associated cosine similarities in abstracts of peer-reviewed papers to explore the trends in scientific literature using alluvial diagrams. Link prediction between entities was conducted on the basis of forecasted values of topological proximity scores. High cosine similarity represents strong relationships between words. We used diachronic word embeddings to capture the semantic shift between various diseases and studied its evolution over time by implementing the Infomap community detection algorithm on networks. Every community or module in the alluvial diagram is ranked by its importance in the network based on the PageRank value of the nodes it contains. The predicted network of February 2021 represented significant themes in the literature, with a large focus on COVID-19 related adverse events and critical conditions, followed by underlying comorbidities that highly influence recovery, symptoms and psychological factors (Supplementary Table 3). The model was used for forecasting inferences for the month of March 2021 based on the trends in node pair proximity measures from the past twelve months (Supplementary Table 4). Our findings based on the model’s predictions suggested that a cluster of autoimmune diseases, multisystem inflammatory syndrome and neurological complications, seen as a part of relatively lower modules in previous months, would assume a higher centrality in March 2021 (Supplementary Figure 6).

Predicting emerging modules of medical entities can also assist in steering research while understanding the themes represented by them. The method presented in this paper has exclusively been applied on networks of diseases and medical conditions and can be extended to various other dynamical analyses such as the evolving impact of entities in dynamic knowledge graphs. A potential advancement in the architecture of temporal link prediction can include larger data and complex models like RNN and LSTM to predict the links (10, 11).

The potential of unsupervised word embeddings and NER was leveraged to extract insights regarding the topmost similar diseases or chemicals with selected keywords. Vaccine, which has been a rising topic lately, had the highest cosine similarity with Ad26.COV2.S (also known as Ad26) and mRNA-1273, which are a few of the most discussed candidate vaccines for COVID19 in the literature. Since the immunization drive has started across the globe, a stern feeling of ‘hesitancy’ has developed towards vaccines. ‘Comorbidity’ is found to have a high similarity with hyperlipidemia, diabetes mellitus, heart as well as kidney diseases. A number of long-term effects have been reported post-recovery due to a weakened immune system. Exploring ‘adverse effects’ as a keyword depicted correlations with retinal adverse events, social-health crisis and adrenal crisis. People with obsessive-compulsive spectrum disorders have been affected adversely as well. ‘Social’ factors were found to have the highest similarity with connectedness and an increase in family violence and psychological damage. It also highlights the existing gap between rural and urban communities. The economic recession caused by the pandemic has also led to loneliness and social anxiety which was captured by the word embeddings as well. ‘Psychological’ health has been negatively impacted due to worry and stress over the coronavirus, which is characterized by the aggravation of conditions such as PTSD and an intuitively high cosine similarity with ‘coping behaviors’. A projection of the top 100 entities nearest to the keyword ‘mental disorders’ in a three-dimensional space of word embeddings reduced using Principal Component Analysis has been shown in Supplementary Figure 4. The presence of alcohol abuse, substance abuse and suicidal behaviors in the same space explains that the Word2Vec model trained on abstracts of vast COVID-19 literature effectively captured latent associations among general keywords and medical entities. Detailed descriptions of the analysis can be found in Table 2.

**Table 2:** Topmost similar entities (diseases, conditions or chemicals) with selected keywords: ‘vaccine’, ‘comorbidity’, ‘adverse effects’, ‘social’ and ‘psychological’, in descending order of cosine similarity calculated using the word embeddings generated from the Word2Vec model trained on the entire corpus. The cosine similarity of words have also been used as a feature for link prediction.

| *vaccine* | *comorbidity* | *adverse_effects* | *social* | *psychological* |
| --- | --- | --- | --- | --- |
| hesitancy | CCI | cognitive and prosocial coping behaviors | social cohesion | distress |
| Ad26 | comorbid | social-health crisis | employment loss | social anxiety |
| MMR | CHF (congestive heart failure) | obsessive-compulsive spectrum disorders | social distancing behaviors | technostress |
| OPV | concomitant diseases | phosphate hydroxychloroquine | connectedness | psychological distress |
| BNT162b2 | ischemic heart disease | hydroxychloroquine-sulphate | self-disclosure | vicarious traumatization |
| mRNA-1273 | hypertension | retinal adverse | family violence | psychopathology |
| ChAdOx1 | chronic renal disease | delays in skin cancer | social connectedness | coping behaviors |
| Calmette-Guérin | cardiovascular comorbidity | Refeeding syndrome | social stigma | maladjustment |
| drug/vaccine | liver cirrhosis | respiratory affliction | psychological damage | post-traumatic stress symptoms |
| Moderna | chronic renal disease | adrenal crisis | compulsive | peritraumatic distress |

An open-source dashboard called *EvidenceFlow*, has been built, which has surfaced as a template for analyzing themes and communities of diseases mentioned in research articles over time. The dashboard also allows the user to unravel the literature with an interactive map of embeddings based on the visualization provided by Tensorboard. The dashboard aims to track literature trends using alluvial diagrams, community detection and projection of influential entities through network analysis across different months. An extension of the current tool opens new directions for analysis of higher frequency and a greater variety of entities such as genes, drugs or adverse drug effects.

## Materials and Methods

### Dataset

The dataset was created using more than 77,000 peer-reviewed publications and more than 11,000 WHO vetted preprints present in the *WHO Database* (2) from February 2020 to February 2021 (Supplementary Figure 1). The frequency of articles with respect to specific categories and keywords have been depicted in Supplementary Table 2.

### Text Pre-processing and Exploratory Data Analysis

Formatting of text and removal of white spaces, punctuation, digits and stop words were carried out on lowercase converted text using NLTK package (12). Word frequency distributions were visualized as chatter plots using the ggplot2 package (13).

### Named Entity Recognition

Named Entity Recognition was used to extract two types of entities: diseases and chemicals, from the original abstracts of vetted research articles using a model pre-trained on BC5CDR corpus by SciSpacy, an open-source project developed for Biomedical Natural Language Processing (14). The model identifies the entities with an F1 score of 84.49%. The words extracted under the category of diseases also contained symptoms, adverse effects, conditions, disorders and syndromes. All of these are collectively referred to as diseases in the further sections. Entities were further used for creating networks to study the trends through alluvial diagrams and for predicting temporal links between nodes across past and upcoming months.

### Unsupervised Word Embeddings

A low-dimensional representation for the words present in the corpus of abstracts was learned using the Word2Vec model with skip-gram algorithm, one-hot encoding and fixed window size of five, implemented in Gensim (15, 16, 17). The word embeddings of the extracted entities were used to calculate the cosine distance between the word vectors in order to analyze the dis(similarity) between entities. Visualization of the word vectors was carried out using TensorFlow Embedding Projector (18) to allow interactive exploration of relationships between diseases and chemicals. Supplementary Figure 1(c) demonstrates a snapshot of entities closest to the keyword ‘COVID-19 disease’. For the creation of each month’s network of entities, separate Word2Vec models were trained in order to allow capturing of semantic shifts in word similarities in the literature published over time.

### Longitudinal Entity Networks and Communities

Weighted networks were constructed using similarity between word vectors of extracted entities as edge weights. A union of all entities with top ten percentile cosine similarity across each month from February 2020 to February 2021 were preserved as nodes in the networks. Community detection was done over the monthly networks using the Infomap algorithm (19). Semantic change in the word embeddings led to the formation of communities which shifted as emerging themes over months. The importance of each node was tracked using an alluvial visualization based on PageRank values (20). Detailed steps with parameters are available in the Supplementary Information.

### Time Series Forecasting of Proximity Scores

In order to predict the existence of links between nodes in temporal networks of subsequent months, we computed five neighborhood proximity scores for the network of each month. Jaccard Similarity (21), Preferential Attachment (22), Adamic Adar Similarity (23), and Common Neighbours (24) were used as topology-based features and Cosine Similarity between the entities represented by the nodes was used as a semantic feature. These proximity scores based upon network topology were calculated using the NetworkX package (25). The range of Adamic Adar Similarity, Common Neighbours, and Preferential Attachment values lies between (0.00 - ∞), while that of Jaccard Similarity and Cosine Similarity lies in the range of (0.00 - 1.00). In order to scale the values, we normalized the former three scores in each network to bring them in the range of (0.00 - 1.00).

For each node pair, every proximity score was modeled as a time series and the value was predicted for the subsequent month using the Auto-Regressive Integrated Moving Average, or the ARIMA model (26). Non-stationary time series were passed through the Random Walk order of the model. For validating, proximity scores for the network at timestamp *τ+1* were predicted based on their respective past values in the networks till timestamp *τ*. The performance of the model was assessed by comparing the predictions with the original proximity scores using the Mean Squared Error (Supplementary Table 5).

### Temporal Link Prediction between Entities

The proximity scores predicted using the ARIMA model were further used to identify the occurrence of a link between entities in network G_*τ*+1_ based on the proximity scores and links in all previous networks [G_1_, G_2_, G_3_, …, G_*τ*_], using supervised machine learning. The ground truth label for the presence of a link in the longitudinal entity networks was defined as 1 if the cosine similarity between entities was greater than the threshold, that is, 90th percentile of the cosine similarities between top-N(100) diseases in that month’s abstracts, otherwise 0.

We implemented the proposed link prediction approach using Logistic Regression (27), Random Forests (28), SVM (29), AdaBoost (30), XGBoost (31). For training the models, proximity scores Jaccard Coefficient, Preferential Attachment, Adamic Adar Index and Common Neighbors were used as features for node pairs in networks from February 2020 till December 2020. Validation of the model was done over the subsequent month of January 2021, taking its predicted proximity scores as features. For comparing the predicted links with the ground truth links, Area Under Receiver Operator Characteristic Curve (AUC ROC) score was evaluated after selecting the optimal threshold for binary classification to deal with imbalanced data. While training, validating and testing the model, we did not use Cosine Similarity as a feature as it was the identifier variable for the link. Testing of the model was performed on the predicted proximity scores of February 2021. Random Forest was found to be the best model based on AUC ROC score of 0.77. The full detail of the algorithm and features are available in the Supplementary Information.

### Community Detection on Predicted Networks

The links between node pairs predicted by the Random Forest model were used to create networks weighted by cosine similarity scores predicted by the ARIMA model. Infomap algorithm was applied on the predicted as well as original test network to cluster the nodes into 10 modules. The modules were compared using Intersection Over Union (IOU) (32) using the following formula:

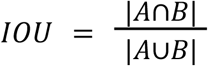

where A represents set of nodes in the predicted i ^th^ module, i ∈ {1, 2, …, 10}, and B represents set of nodes in the original j^th^ module, j ∈ {1, 2, …, 10}.

## Data Availability

All the data used in this study are publicly available from the WHO Covid-19 Global Literature on coronavirus disease maintained at https://www.who.int/emergencies/diseases/novel-coronavirus-2019/global-research-on-novel-coronavirus-2019-ncov. Our analysis and the interactive resource EvidenceFlow is publicly available in a user-friendly fashion at https://evidenceflow.tavlab.iiitd.edu.in/

## Acknowledgments

This work was partially supported by the Wellcome Trust/DBT India Alliance Fellowship IA/CPHE/14/1/501504 awarded to Tavpritesh Sethi. We also acknowledge support from the Center of Excellence in Healthcare and the Center of Excellence in Artificial Intelligence at IIIT-Delhi.

