## Supplementary Material for "Predicting Emerging Themes in Rapidly Expanding COVID-19 Literature with Dynamic Word Embedding Networks and Machine Learning"

### Supplementary Information

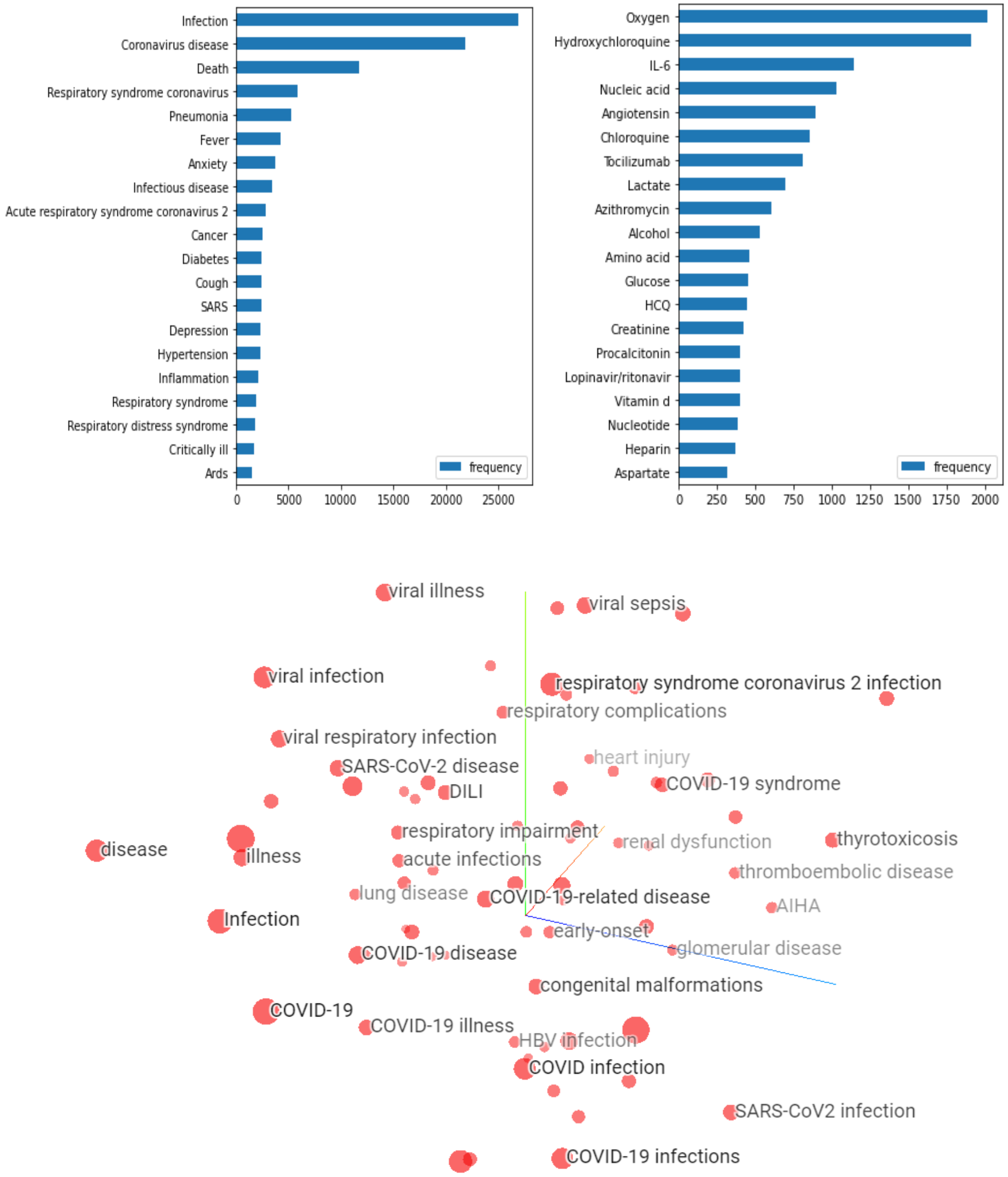


*Supplementary Figure 1: (a) Bar plot (left) showing the frequency of top diseases in the corpus of abstracts extracted using Named Entity Recognition. (b)Bar plot (right) showing the frequency of top chemicals in the corpus of abstracts extracted using Named Entity Recognition. (c) Latent space of word embeddings of diseases and chemicals visualized around the keyword 'COVID-19 disease', displaying 100 isolated points nearest to it.*

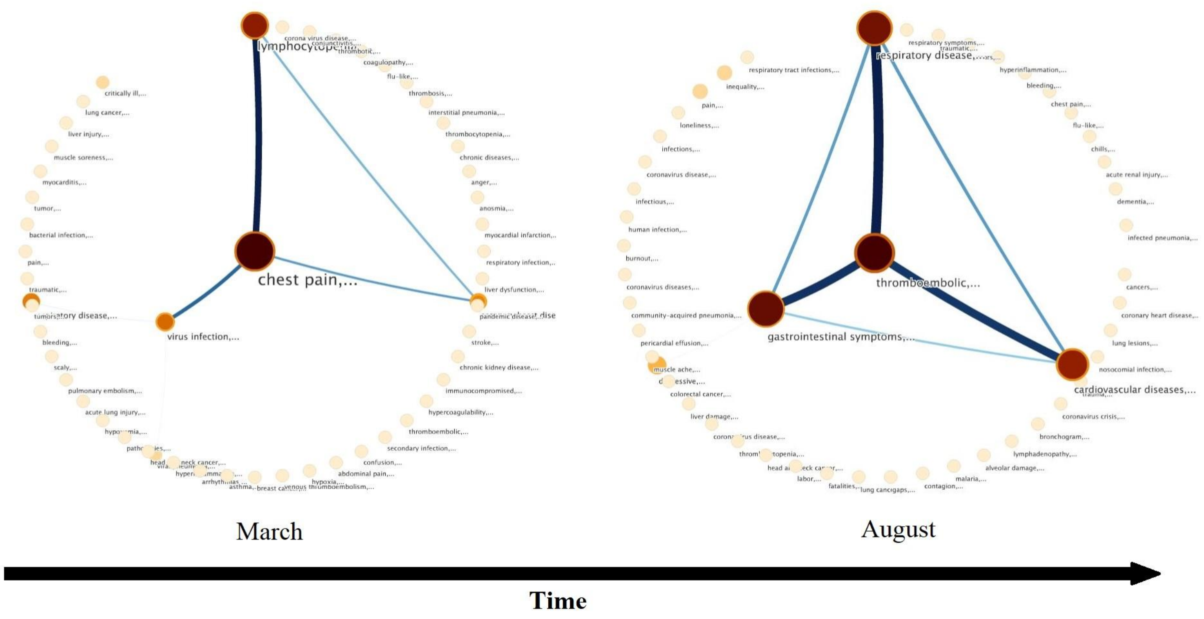


*Supplementary Figure 2: Longitudinal Community network from March 2020 to August 2020. The emergence of Thromboembolic complications as a major theme by August 2020 has been illustrated in these networks.*

| Model | AUCROC | Accuracy | F1 score |
|---|---|---|---|
| **Logistic Regression** | 0.720 ± 0.0014 | 0.78 ± 0.0007 | 0.53 ± 0.0016 |
| **Random Forest** | **0.77 ± 0.0012** | **0.78 ± 0.0008** | **0.53 ± 0.0017** |
| **Support Vector Machine** | 0.70 ± 0.0013 | 0.76 ± 0.0008 | 0.52 ± 0.0017 |
| **ADABoost** | 0.71 ± 0.0014 | 0.77 ± 0.0008 | 0.52 ± 0.0017 |
| **XGBoost** | 0.74 ± 0.0011 | 0.78 ± 0.0008 | 0.51 ± 0.0017 |

*Supplementary Table 1: Results of temporal link prediction between entities for the month of February 2021, with a margin of error for 95% Confidence Intervals. The mean value of metrics has been recorded by testing the models on a resampled test set.*

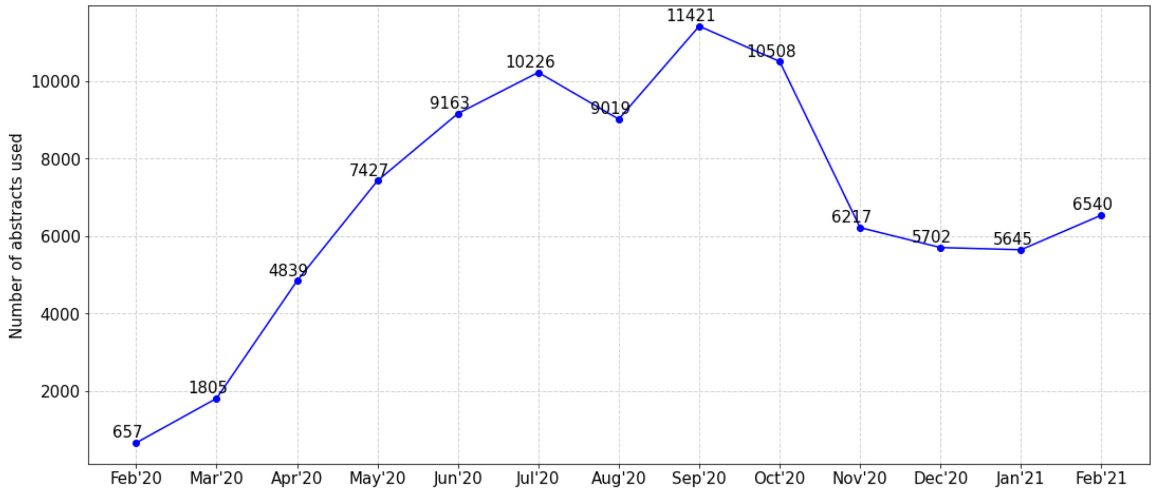


*Supplementary Figure 3: Showing the number of articles occurring each month. The curve depicts that there has been an increase in the number of articles across each month since February 2020.*

| **Category** | **Keywords** | **Article Count** |
|---|---|---|
| Respiratory Disease | Pneumonia<br>Cardio<br>Rheumatic | 2201<br>992<br>149 |
| Infectious Disease | HIV<br>Diarrhea<br>Tuberculosis | 362<br>61<br>103 |
| Maternal & Child Health | Pregnant<br>Maternal health<br>Child | 422<br>8<br>1832 |
| Cancer | Cancer | 1511 |
| Risk | Risk | 3370 |

*Supplementary Table 2: Frequency of articles belonging to specific categories. The counts are based on the occurrence of the given keywords in the article's title.*

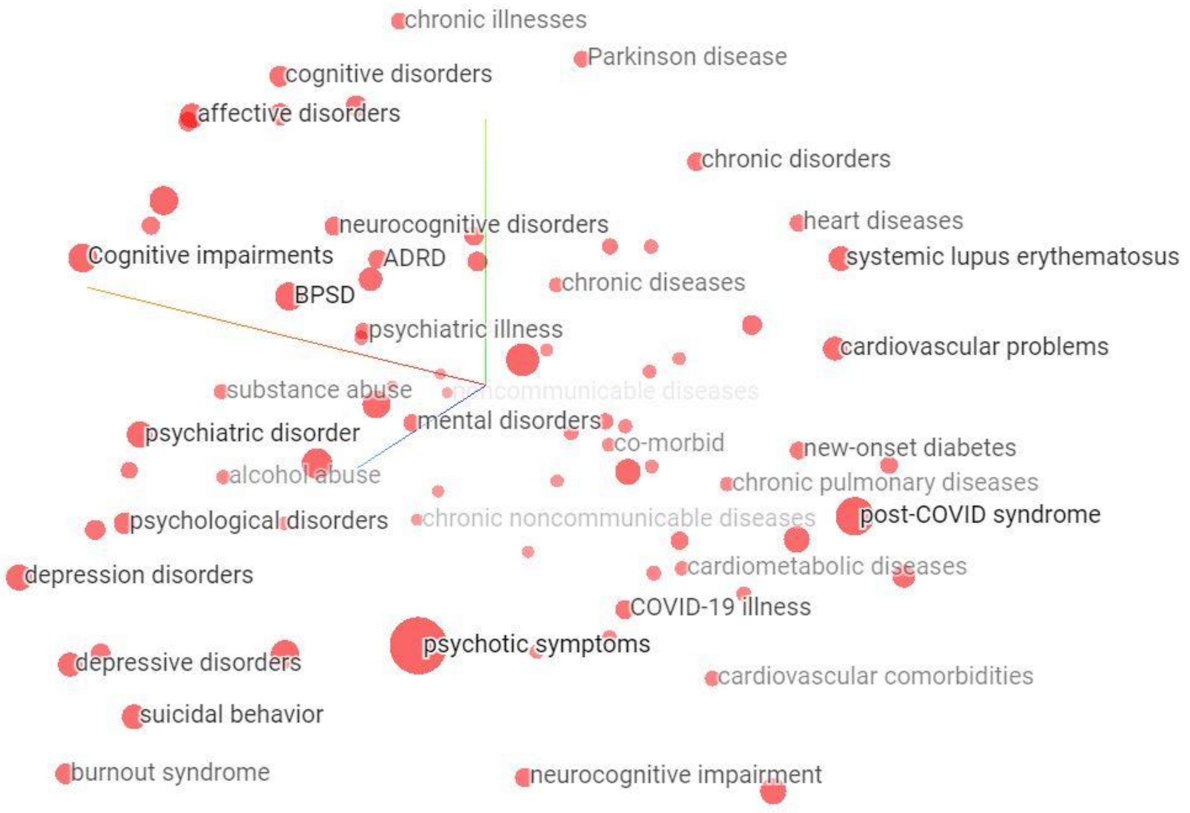


*Supplementary Figure 4: Latent space of word embeddings of diseases and chemicals visualized around the keyword 'mental disorders', displaying 100 isolated points nearest to it.*

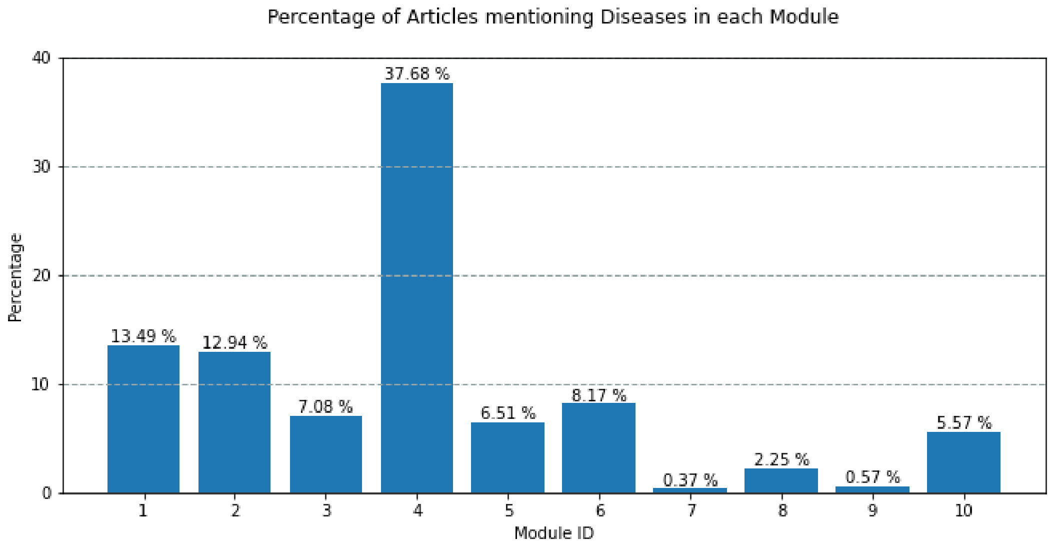


*Supplementary Figure 5: Percentage of abstracts of articles published in February 2021 mentioning diseases belonging to each module in predicted network.*

| | **Theme: Subset of nodes from different modules** |
|---|---|
| Module 1 | Adverse events:<br>respiratory failure, pulmonary embolism, acute kidney injury, hyperinflammation, sepsis |
| Module 2 | Comorbidities:<br>obesity, breast cancer, diabetes, hypertension, asthma, dementia, chronic kidney disease |
| Module 3 | Symptoms:<br>anosmia, shortness of breath, fever, fatigue, diarrhea, myalgia, dyspnea, headache, cough |
| Module 4 | Respiratory illness:<br>MERS, acute respiratory syndrome coronavirus 2, respiratory infections |
| Module 5 | Viral disease:<br>toxicity, viral infection, tuberculosis, malaria |
| Module 6 | Psychological conditions:<br>PTSD, psychological distress, loneliness, burnout, anxiety, depression, traumatic |
| Module 7 | opacities |
| Module 8 | fracture, trauma, stroke, injuries |
| Module 9 | panic, inequality |
| Module 10 | infections, fatalities |

*Supplementary Table 3: Results of community detection from predicted network for February 2021. A subset of nodes is mentioned which broadly signifies a theme for the given module.*

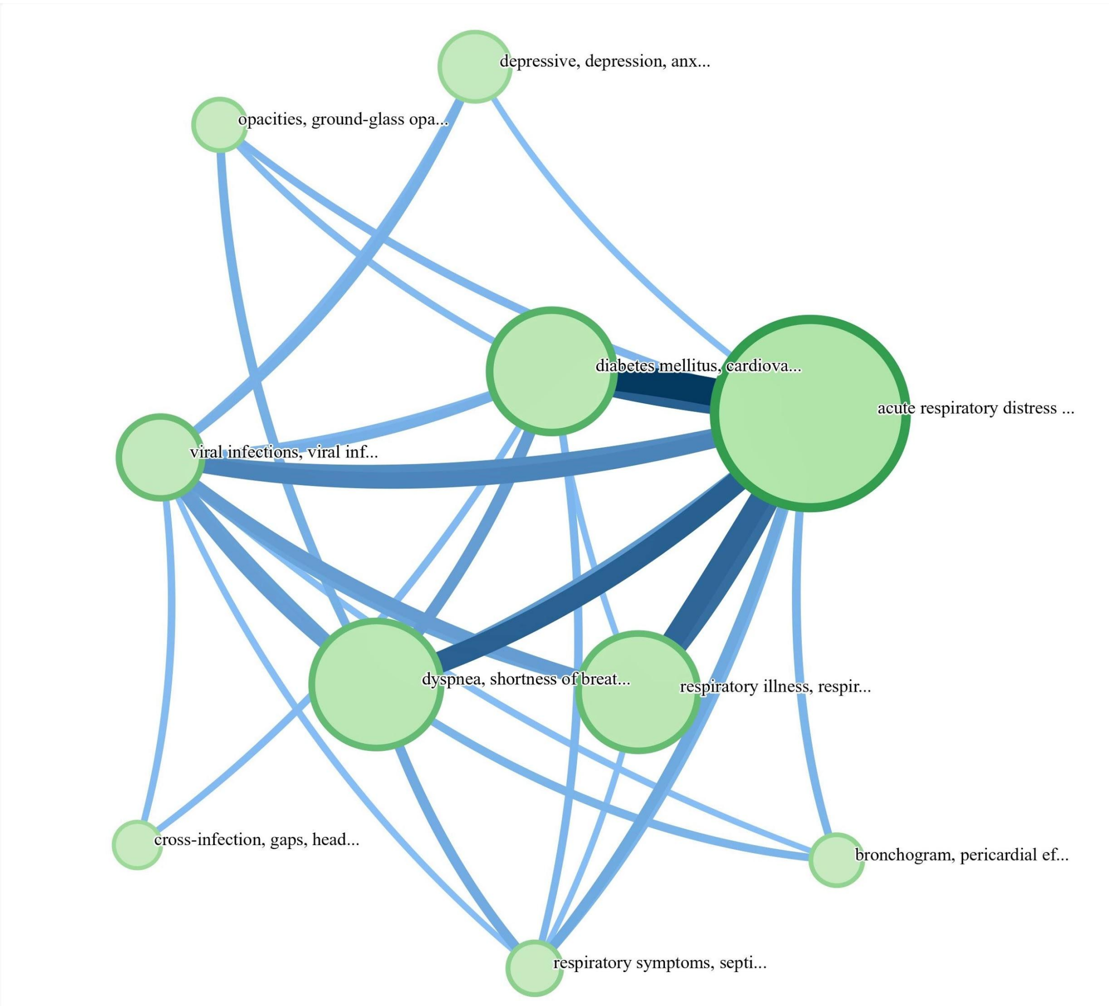


*Supplementary Figure 6: Multi-level network generated from Infomap illustrating the links between different communities for the month of March, 2021.*

| | **Theme: Subset of Intersecting nodes** |
|---|---|
| Module 1 | Adverse events:<br>ARDS, myocarditis, multisystem inflammatory syndrome, autoimmune diseases, ischemic stroke, arrhythmia, hypercoagulability, neurological symptoms |
| Module 2 | Symptoms:<br>Dyspnea, shortness of breath, headache, gastrointestinal symptoms, vomiting, myalgia, diarrhea, anosmia |
| Module 3 | Comorbidities:<br>diabetes mellitus, cardiovascular disease, chronic obstructive pulmonary disease, asthma, hypertension, chronic kidney disease |
| Module 4 | Respiratory illness: |

| | |
|---|---|
| | respiratory illness, respiratory infections, respiratory disease, acute respiratory syndrome, mers |
| Module 5 | Infections & Cancer:<br>viral infections, confusion, scaly, panic, bacterial infection, malignancy, toxicity, cancer |
| Module 6 | Psychological conditions:<br>depressive, depression, anxiety, psychological distress, burnout, loneliness, insomnia |
| Module 7 | respiratory symptoms, septic shock, co-infection, respiratory tract infection, liver dysfunction, lymphadenopathy, muscle soreness, thrombocytopenia |
| Module 8 | Bronchogram, pericardial effusion, chest pain, bronchiectasis, coronary heart disease |
| Module 9 | cross-infection, gaps, head and neck cancer, nosocomial infection, 'labor |
| Module 10 | interstitial pneumonia, conjunctivitis |

*Supplementary Table 4: Results of community detection from predicted network for March 2021. A subset of intersecting nodes is mentioned which collectively signify a theme.*

**Longitudinal Entity Networks and Communities.**

Based on the frequency, we took into account the top N(100) entities from the abstracts of papers published in each month. Corresponding networks were constructed using the following algorithm:

1. For every month $\tau$ in T months,
2. Top N entities extracted from the corpus of abstracts using NER.
3. All possible pairs of entities (${}^{N}C_2$ combinations) created to represent node pairs in a network. The weight of the edges is valued by the Cosine Similarity between the entities generated from the Word2Vec model trained on the corpus of month $\tau$.
4. Edges weighing more than the 90th percentile of the weight are preserved.
5. A union of node pairs ($\Delta = \bigcup(u,v)$) from all the months is taken in order to have a common set of nodes in each. Every month $\tau$ is depicted by a network $G_{\tau}$.

**Temporal Link Prediction between Entities.**

The algorithm used to predict links in the network at timestamp $\tau$+1 is demonstrated below.

1. For each node pair *(u,v)* $\in \Delta$ in $G_t$, t $\in$ {1, 2,…,$\tau$), five proximity scores were calculated based on the topological features of the graph and semantic similarity between entities.
   - Cosine Similarity, from the Word2vec model trained on the corpus of month $\tau$.
   - Jaccard Coefficient (JC)
   - Number of Common Neighbors (CN)
   - Preferential Attachment (PA)
   - Adamic-Adar Index (AA)

   Since the range of CN, PA and AA lies between 0.00 − ∞, we normalized the respective scores in the range of 0.00 − 1.00 in each network $G_t$.
2. For each proximity score, a ($|\Delta| \times \tau$) matrix was created where $|\Delta|$represents the number of node pairs and $\tau$ represents the number of months taken in the training set. This matrix stores the value of the proximity score for node pair *(u,v)* $\in \Delta$ at timestamp t.
3. For each node pair, the value of proximity score is forecasted at timestamp $\tau$+1 using the ARIMA model (p=1, d=0, q=0) if the series is stationary, else random walk order is used (p=0, d=1, q=0). Mean Squared Error in the predictions of each proximity score for the month of January 2021 and February 2021 are shown in Supplementary Table 3.
4. Training of the classification model is done using four topological proximity scores as features (excluding cosine similarity as it is an identifier variable) from the networks $G_1$, $G_2$,…, $G_{\tau.}$
5. Testing set features represent the four predicted proximity scores $G_{\tau+1}$ for all the node pairs *(u,v)* $\in \Delta$.
6. Due to a high imbalance between positive and negative labels, the Receiver Operator Curve (ROC) was used to obtain an optimal threshold for the binary classification, using Youden's J Statistic in the following formula:

   $$J = TPR - FPR$$

   where, TPR = True Positive Rate and FPR=False Positive Rate.

J represents an array of differences between TPR and FPR of different points on the ROC curve. The index of the maximum value of J, that is, argmax(J), is used as a criterion for selecting the point which may represent the optimum threshold.

7. The optimum threshold was used to binarize the predicted probabilities into 0 and 1.
8. Links predicted from the model were verified across the ground truth links of $G_{\tau+1}$. The average performance metrics of the model was obtained by resampling the test set repeatedly for 100 times and testing 1000 samples in each iteration. The margin of error for 95% confidence intervals was also calculated.

| **Proximity Score** | **MSE** |
|---|---|
| Cosine Similarity | 0.113 |
| Jaccard Coefficient | 0.029 |
| Number of Common Neighbors | 0.030 |
| Preferential Attachment | 0.043 |
| Adamic-Adar Index | 0.031 |

| **Proximity Score** | **MSE** |
|---|---|
| Cosine Similarity | 0.102 |
| Jaccard Coefficient | 0.045 |
| Number of Common Neighbors | 0.056 |
| Preferential Attachment | 0.044 |
| Adamic-Adar Index | 0.058 |

*Supplementary Table 5: Mean Squared Error (MSE) between original and predicted proximity scores for the network of (a) January 2021 (b) February 2021.*
